# Protection conferred by vaccine plus previous infection (hybrid immunity) with vaccines of three different platforms during the Omicron variant period in Brazil

**DOI:** 10.1101/2022.04.12.22273752

**Authors:** Thiago Cerqueira-Silva, Vinicius de Araujo Oliveira, Enny S. Paixão, Pilar Tavares Veras Florentino, Gerson O. Penna, Neil Pearce, Guilherme L. Werneck, Maurício L. Barreto, Viviane S. Boaventura, Manoel Barral-Netto

## Abstract

Hybrid immunity (infection plus vaccination) provided high protection against infection and severe disease in the periods of delta and gamma variants of concern. However, the protection of hybrid immunity in the Omicron era remains unknown. We performed a test-negative study using Brazilian national databases between January 01 and March 22, 2022, a period of predominant circulation of the Omicron variant in Brazil. Hybrid immunity offered low protection against infection, with rapid waning, compared to unvaccinated with or without previous infection. For severe illness (hospitalisation or death), the protection, although already high for unvaccinated pre-infected increased regardless of the type of vaccine (Ad26.COV2.S, BNT162b2, ChAdOx-1 or CoronaVac).

In conclusion, during the Omicron-dominant period in Brazil, hybrid immunity offered high protection against severe illness and low protection against infection.

---

As of April 7, 2022, it has been estimated that 495 million individuals have been infected by SARS-CoV-2, and at least 11 billion COVID-19 vaccine doses have been administered worldwide^1^. Therefore, understanding hybrid immunity (infection plus vaccination) is crucial to guide future vaccination policies. We have demonstrated that vaccination offered additional protection to that induced by an infection during the Gamma and Delta variants waves in Brazil ^2^. With the emergence of the Omicron variant, vaccine effectiveness (VE) appears to decay^3,4^, but the protection in previously infected vaccinees remains unknown. Here, we analyzed the impact of hybrid immunity in preventing infection and severe outcomes during the circulation of the Omicron variant in Brazil.

Using national databases, we performed a test-negative case-control study as previously described^2^. Cases and controls were defined as individuals with RT-PCR/Lateral-flow tests positive or negative, respectively, between January 01 and March 22, 2022, a period of predominant circulation of the Omicron variant in Brazil (Appendix:pg=3-4). Severe outcomes (hospitalisation or death) were defined as: a positive test obtained from 14 days before to 3 days after hospital admission; death occurring within 28 days after a positive test. We analyzed VE in previously infected vaccines using two references groups: unvaccinated with or without pre-infection. Detailed methods are in the Appendix (page 2).

A total of 918,219 tests (899,050[97.9%] individuals) were included, 476,901 (51.9%) cases, and 441,318 (48.1%) controls, and 323,704 (35.2%) were unvaccinated (22,935[2.4%] with and 300,769 [32.8%] without pre-infection) (Appendix:pg 4-6). Compared to those unvaccinated without pre-infection, the effectiveness of the previous infection in preventing reinfection during the Omicron period was low (28.9%, 95% confidence interval [CI]26.9-30.9), increasing with vaccination with any vaccine type (Ad26.COV2.S, BNT162b2, ChAdOx-1 or CoronaVac), especially after the booster, although this protection waned over time. Protection against severe outcomes after a previous infection was relatively high (85.6%, 95%CI:82.7-88.0), increasing with vaccination (VE ranging from 88.0 to 100%). Compared to those unvaccinated with a previous infection, hybrid immunity showed a modest increase in protection against symptomatic infection, once again waning over time, and substantial protection against severe outcomes after the booster (Figure 1/Appendix:pg=7-8). Similar results were obtained using a matched design (Appendix:pg=9-11).

**Figure 1:**
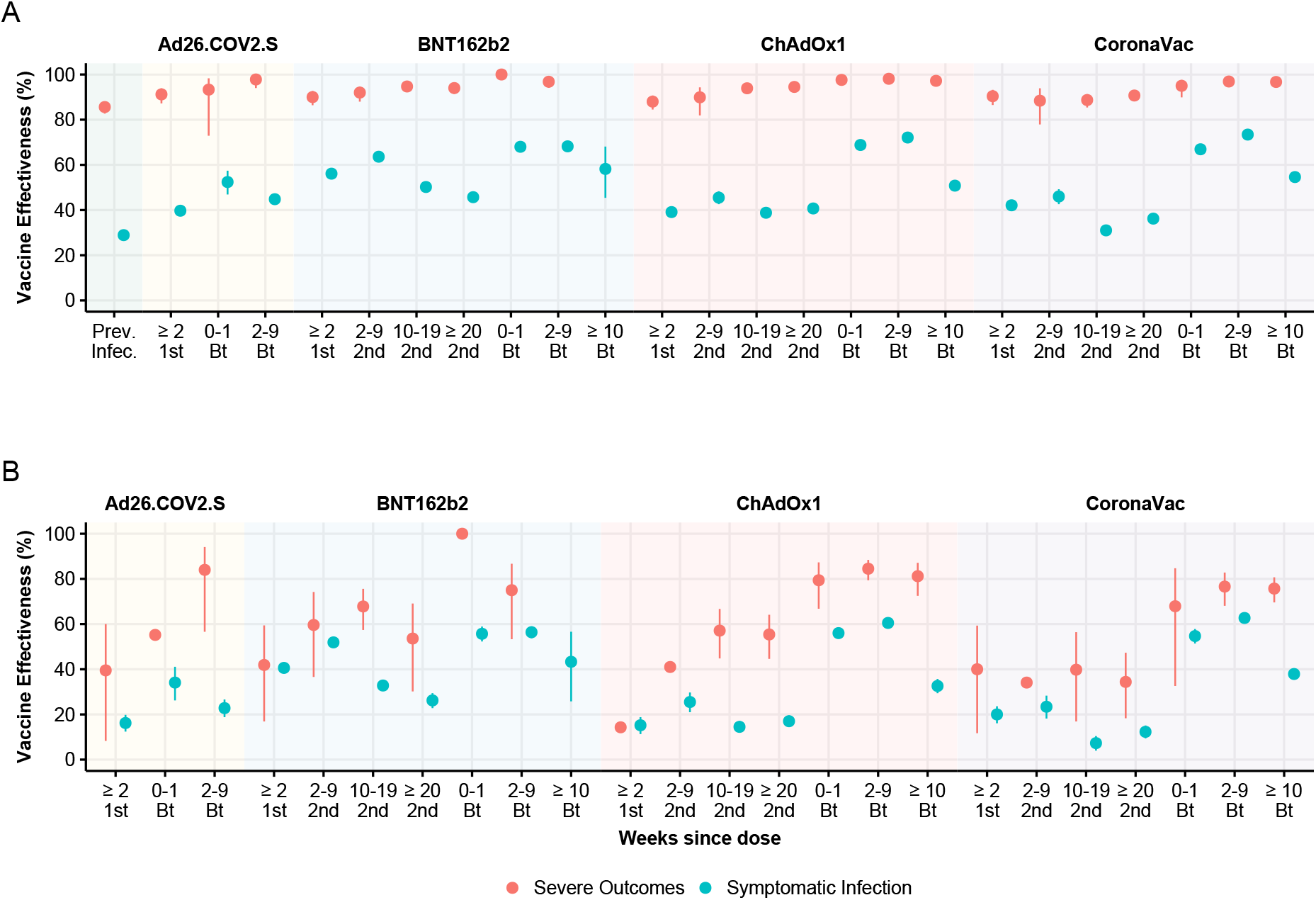
Effectiveness of hybrid immunity against SARS-CoV-2 symptomatic infection and severe outcomes. A) Effectiveness of previous infection and hybrid immunity compared to unvaccinated individuals without previous. B) Effectiveness of hybrid immunity compared to unvaccinated individuals with the previous infection. 1st= First dose, 2nd= Second dose, Bt= Booster dose. To ensure reasonable precision, estimates are shown when there were at least 20 cases or 1000 controls for symptomatic infection, and 10 cases or 500 controls for severe outcomes.

In conclusion, during the Omicron dominant period in Brazil, robust protection against severe disease was offered by a previous infection and this was increased with hybrid immunity (infection+vaccination). However, against symptomatic infection, even boosted individuals with hybrid immunity had lower levels of protection and these waned over time. Booster doses in previously infected individuals offered a moderate but transient gain in protection against symptomatic infection and a slight improvement against severe outcomes.

## Data Availability

One of the study coordinators (MB-N) signed a term of responsibility on using each database made available by the Ministry of Health (MoH). Each member of the research team signed a term of confidentiality before accessing the data. Data was manipulated in a secure computing environment, ensuring protection against data leakage. The Brazilian National Commission in Research Ethics approved the research protocol (CONEP approval number 4.921.308). Our agreement with MoH for accessing the databases patently denies authorization of access to a third party. Any information for assessing the databases must be addressed to the Brazilian MoH at https://datasus.saude.gov.br/, and requests can be addressed to. Herein we used anonymized secondary data following the Brazilian Personal Data Protection General Law (LGPD), but it is vulnerable to re-identification by third parties, as they contain dates of relevant health events regarding the same person. To protect the research participants' privacy, the approved Research Protocol (CONEP approval number 4.921.308) authorizes only the dissemination of aggregated data, such as the data presented here.

