## Appendix for "Protection conferred by vaccine plus previous infection (hybrid immunity) with vaccines of three different platforms during the Omicron variant period in Brazil"

### METHODS

We used a test-negative case control design. We analysed a deterministically linked dataset comprised of the Programa Nacional de Imunizações, which holds records of all vaccines administered in Brazil (BNT162b2, ChAdOx1, Ad26.COV2.S or CoronaVac); the e-SUS Notifica, which contains records of suspected and confirmed COVID-19 in outpatient clinics; and the Sistema de Informação da Vigilância Epidemiológica da Gripe, which holds records of all COVID-19 hospitalisations and deaths. All data were pseudo-anonymized, with a common unique identifier provided by the Brazilian Ministry of Health. The research protocol was approved by the Brazilian National Commission in Research Ethics (CONEP) (approval no. 4.921.308).

All individuals aged 18 years or older who reported COVID-19-like symptoms and were tested for SARS-CoV-2 between January 01 2022, and March 22 2022, were eligible for the study. We excluded: (1) individuals who received a different vaccine for the second dose from the first; (2) individuals whose time interval between the first and second doses was fewer than 14 days; (3) tests with missing information of age, sex, municipality of residence or sample collection date; (4) negative test within 14 days of a previous negative test, as those likely represent the same event; (5) negative test followed by a positive test up to 7 days, likely these were false negatives; (6) any test after a positive test up to 90 days; (7) individuals with a booster dose less than 115 days in those who received CoronaVac, ChAdOx1 and BNT162b2 as primary series; (8) Individuals with more than 3 COVID-19 vaccine doses; (9) Individuals with booster dose different than BNT162b2 in those who received CoronaVac, ChAdOx1 and BNT162b2 as primary series and booster different than Ad26.COV2.S in those who received Ad26.COV2.S as primary series.

Cases of confirmed infection were defined as adults with a positive SARS-CoV-2 RT–PCR or Lateral-flow test and controls with a negative SARS-CoV-2 RT–PCR or Lateral-flow test, both from a sample collected within ten days of symptom onset. Hospital admission up to 14 days after a positive test or a positive test within <72 hours of hospital admission. Mortality outcome was defined as death occurring within 28 days after a positive test.

The odds ratio (OR), comparing the odds of vaccination in persons with symptomatic cases of SARS-CoV-2 infection with those in symptomatic persons who tested negative for SARS-CoV-2, and its associated 95% Confidence Interval (CI) was calculated using binomial logistic regression, adjusting for potential confounders (age (five-year band), sex, calendar week, comorbidities (diabetes mellitus, obesity, chronic kidney disease, cardiac disease, chronic respiratory disease, immunosuppression). The comorbidities were categorized as none, one, two, and at least three in the model. In the model comparing only previously infected individuals, a term indicating time since the first infection was included (categorised as 90-179 days,180-365 days and >365 days). Additional sensitivity analysis was conducted categorized previously infected individuals by time since the first infection and compared them against naive unvaccinated. We also performed a matched design to control for a different vaccination program in each municipality; we matched each case to up two controls (with replacement) by date of testing (±10 days), age (in 5-year bands), municipality of residence (n=5,570 municipalities) and sex, additional terms included in the conditional logistic regression were the number of comorbidities and time since the first infection. All data processing and analyses were performed in R (version 4.1.2)

Supplementary Figure 1: Monthly prevalence of SARS-CoV-2 variants in Brazil among genotyped isolates in the GISAID (global initiative on sharing avian influenza data, accessed April 08, 2022)

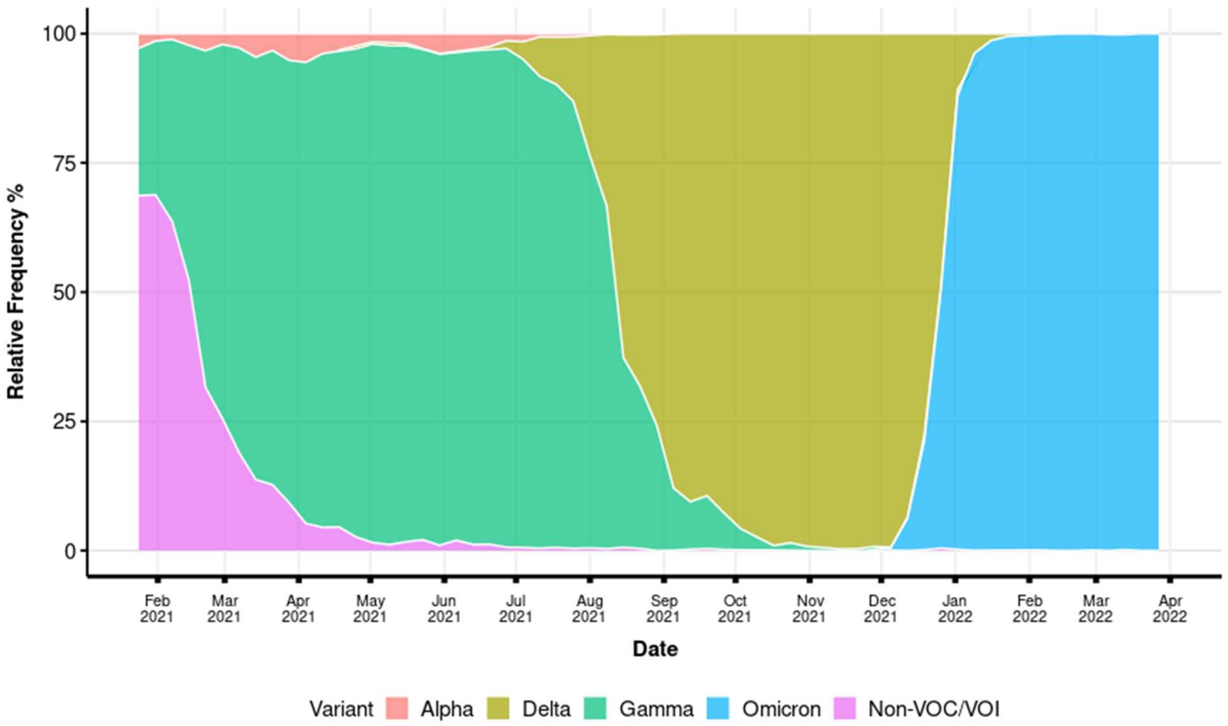

Supplementary Figure 2: Flowchart of the study population from surveillance databases and selection of cases and controls.

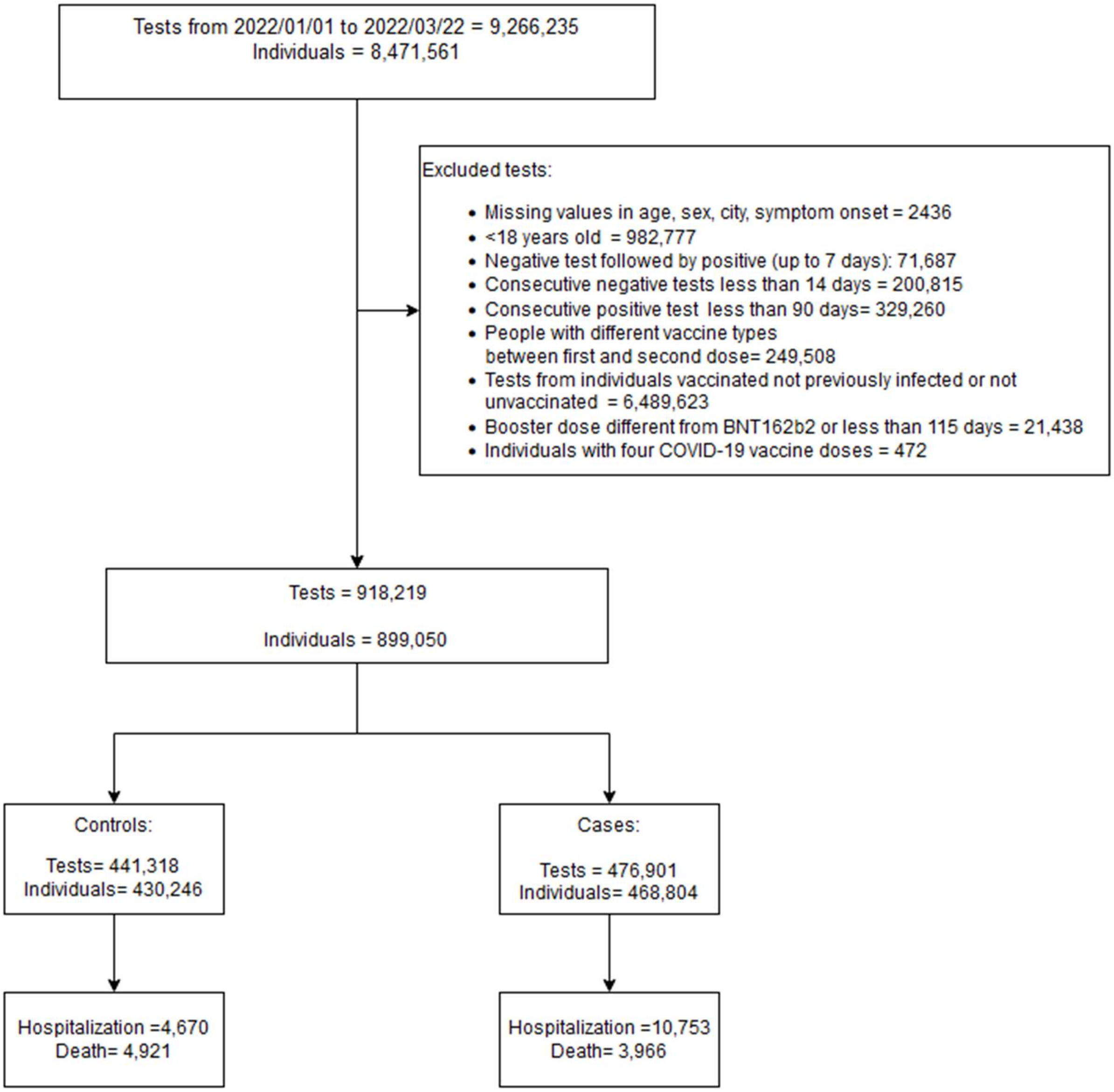

Supplementary Table 1: Characteristics of individuals tested for SARS-CoV-2 in Brazil during the Omicron period, by case/control status

| Characteristic | Cases, N = 476,901 | Controls, N = 441,318 | Overall, N = 918,219 |
|---|---|---|---|
| **No. individuals** | 468,804 (98.3%) | 430,246 (97.5%) | 899,050 (97.9%) |
| **Age median (IQR)-years** | 37 (29, 48) | 36 (27, 48) | 37 (28, 48) |
| **Age Group** | | | |
| 18-64 | 448,432 (94.0%) | 414,979 (94.0%) | 863,411 (94.0%) |
| ≥ 65 | 28,469 (6.0%) | 26,339 (6.0%) | 54,808 (6.0%) |
| **Sex-Female** | 266,191 (55.8%) | 255,818 (58.0%) | 522,009 (56.9%) |
| **Race** | | | |
| White | 241,377 (50.6%) | 230,469 (52.2%) | 471,846 (51.4%) |
| Black | 18,398 (3.9%) | 19,324 (4.4%) | 37,722 (4.1%) |
| Asian | 9,100 (1.9%) | 8,481 (1.9%) | 17,581 (1.9%) |
| Mixed | 156,936 (32.9%) | 148,269 (33.6%) | 305,205 (33.2%) |
| Indigenous | 706 (0.1%) | 832 (0.2%) | 1,538 (0.2%) |
| (Missing) | 50,384 (10.6%) | 33,943 (7.7%) | 84,327 (9.2%) |
| **Previous SARS-CoV-2 Infection** | | | |
| No | 190,173 (39.9%) | 110,596 (25.1%) | 300,769 (32.8%) |
| 90-179d | 13,772 (2.9%) | 23,730 (5.4%) | 37,502 (4.1%) |
| 180-365d | 147,791 (31.0%) | 179,735 (40.7%) | 327,526 (35.7%) |
| 366+d | 125,165 (26.2%) | 127,257 (28.8%) | 252,422 (27.5%) |
| **Test type** | | | |
| Lateral-flow | 375,645 (78.8%) | 373,276 (84.6%) | 748,921 (81.6%) |
| RT-PCR | 101,256 (21.2%) | 68,042 (15.4%) | 169,298 (18.4%) |
| **Residence in capital state** | 89,997 (18.9%) | 92,278 (20.9%) | 182,275 (19.9%) |
| **Macroregion** | | | |
| North | 31,977 (6.7%) | 23,902 (5.4%) | 55,879 (6.1%) |
| Northeast | 69,955 (14.7%) | 63,735 (14.4%) | 133,690 (14.6%) |
| Southeast | 192,936 (40.5%) | 181,589 (41.1%) | 374,525 (40.8%) |
| South | 131,345 (27.5%) | 126,547 (28.7%) | 257,892 (28.1%) |
| Central-west | 50,688 (10.6%) | 45,545 (10.3%) | 96,233 (10.5%) |
| **Municipality deprivation quintile** | | | |
| 1 (Least) | 157,296 (33.0%) | 158,773 (36.0%) | 316,069 (34.4%) |
| 2 | 89,553 (18.8%) | 82,398 (18.7%) | 171,951 (18.7%) |
| 3 | 91,750 (19.2%) | 84,916 (19.2%) | 176,666 (19.2%) |
| 4 | 88,074 (18.5%) | 71,223 (16.1%) | 159,297 (17.3%) |
| 5 (Most) | 50,059 (10.5%) | 43,913 (10.0%) | 93,972 (10.2%) |
| (Missing) | 169 (0.0%) | 95 (0.0%) | 264 (0.0%) |
| **Diabetes Mellitus** | 15,411 (3.2%) | 17,922 (4.1%) | 33,333 (3.6%) |
| **Obesity** | 7,246 (1.5%) | 8,267 (1.9%) | 15,513 (1.7%) |
| **Immunosuppression** | 3,594 (0.8%) | 3,953 (0.9%) | 7,547 (0.8%) |
| **Chronic respiratory disease** | 14,890 (3.1%) | 18,555 (4.2%) | 33,445 (3.6%) |
| **Chronic cardiac disease** | 29,036 (6.1%) | 33,348 (7.6%) | 62,384 (6.8%) |
| **Chronic Kidney disease** | 1,957 (0.4%) | 1,779 (0.4%) | 3,736 (0.4%) |
| **Number of medical comorbidities** | | | |
| 0 | 420,220 (88.1%) | 376,597 (85.3%) | 796,817 (86.8%) |
| 1 | 43,942 (9.2%) | 49,133 (11.1%) | 93,075 (10.1%) |
| 2 | 10,398 (2.2%) | 12,597 (2.9%) | 22,995 (2.5%) |
| ≥3 | 2,341 (0.5%) | 2,991 (0.7%) | 5,332 (0.6%) |
| **Hospitalization** | 10,753 (2.3%) | 4,670 (1.1%) | 15,423 (1.7%) |
| **Death** | 3,966 (0.8%) | 980 (0.2%) | 4,946 (0.5%) |
| **Severe outcomes** | 11,467 (2.4%) | 4,921 (1.1%) | 16,388 (1.8%) |

Supplementary Table 2: Vaccination Status of individuals tested for SARS-CoV-2 in Brazil, according to the test result and severity

| Status | Controls, N = 441,318 | Symptomatic Infection, N = 465,434 | Severe Outcomes, N = 11,467 |
|---|---|---|---|
| **Unvaccinated without previous infection** | 110,596 (36.8%) | 180,583 (60.0%) | 9,590 (3.2%) |
| **Individuals with previous infection** | | | |
| **Unvaccinated** | 10,332 (45.0%) | 12,459 (54.3%) | 144 (0.6%) |
| **Ad26.COV2.S** | | | |
| First dose 0-1 wk | 21 (63.6%) | 12 (36.4%) | 0 (0.0%) |
| First dose ≥2 wk | 6,565 (48.2%) | 7,017 (51.6%) | 29 (0.2%) |
| Second dose 0-1 wk | 685 (51.9%) | 634 (48.0%) | 2 (0.2%) |
| Second dose 2-9 wk | 4,370 (49.4%) | 4,479 (50.6%) | 4 (0.0%) |
| Second dose 10-19 wk | 466 (69.4%) | 204 (30.4%) | 1 (0.1%) |
| Second dose ≥20 wk | 1 (14.3%) | 6 (85.7%) | 0 (0.0%) |
| **BNT162b2** | | | |
| First dose 0-1 wk | 206 (65.4%) | 108 (34.3%) | 1 (0.3%) |
| First dose ≥2 wk | 11,131 (58.4%) | 7,876 (41.3%) | 43 (0.2%) |
| Second dose 0-1 wk | 639 (67.0%) | 314 (32.9%) | 1 (0.1%) |
| Second dose 2-9 wk | 13,187 (63.4%) | 7,593 (36.5%) | 24 (0.1%) |
| Second dose 10-19 wk | 50,639 (53.5%) | 43,839 (46.4%) | 93 (0.1%) |
| Second dose ≥20 wk | 8,130 (58.4%) | 5,760 (41.4%) | 30 (0.2%) |
| Booster dose 0-1 wk | 2,245 (62.2%) | 1,362 (37.8%) | 0 (0.0%) |
| Booster dose 2-9 wk | 4,692 (69.7%) | 2,025 (30.1%) | 11 (0.2%) |
| Booster dose ≥10 wk | 274 (78.3%) | 76 (21.7%) | 0 (0.0%) |
| **ChAdOx1** | | | |
| First dose 0-1 wk | 22 (56.4%) | 16 (41.0%) | 1 (2.6%) |
| First dose ≥2 wk | 6,506 (48.7%) | 6,776 (50.8%) | 69 (0.5%) |
| Second dose 0-1 wk | 189 (56.9%) | 143 (43.1%) | 0 (0.0%) |
| Second dose 2-9 wk | 3,149 (51.9%) | 2,908 (47.9%) | 12 (0.2%) |
| Second dose 10-19 wk | 37,276 (46.4%) | 42,941 (53.4%) | 137 (0.2%) |
| Second dose ≥20 wk | 35,767 (50.2%) | 35,200 (49.4%) | 322 (0.5%) |
| Booster dose 0-1 wk | 6,840 (62.5%) | 4,086 (37.3%) | 20 (0.2%) |
| Booster dose 2-9 wk | 25,328 (68.3%) | 11,668 (31.5%) | 86 (0.2%) |
| Booster dose ≥10 wk | 7,663 (59.5%) | 5,182 (40.2%) | 40 (0.3%) |
| **CoronaVac** | | | |
| First dose 0-1 wk | 87 (52.4%) | 79 (47.6%) | 0 (0.0%) |
| First dose ≥2 wk | 5,584 (51.3%) | 5,254 (48.3%) | 37 (0.3%) |
| Second dose 0-1 wk | 243 (55.4%) | 196 (44.6%) | 0 (0.0%) |
| Second dose 2-9 wk | 2,325 (52.5%) | 2,096 (47.3%) | 10 (0.2%) |
| Second dose 10-19 wk | 18,857 (47.9%) | 20,435 (51.9%) | 62 (0.2%) |
| Second dose ≥20 wk | 24,452 (48.2%) | 25,913 (51.1%) | 326 (0.6%) |
| Booster dose 0-1 wk | 2,426 (61.9%) | 1,486 (37.9%) | 8 (0.2%) |
| Booster dose 2-9 wk | 11,492 (69.0%) | 5,094 (30.6%) | 70 (0.4%) |
| Booster dose ≥10 wk | 28,933 (56.9%) | 21,614 (42.5%) | 294 (0.6%) |

Supplementary Table 3. Effectiveness of hybrid immunity against SARS-CoV-2 symptomatic infection and severe outcomes, compared to individuals unvaccinated without previous infection and those unvaccinated with the previous infection.

| | VE% (95% CI) | | | |
|---|---|---|---|---|
| **Status** | **Symptomatic Infection** | **Severe Outcomes** | **Symptomatic Infection** | **Severe Outcomes** |
| **Unvaccinated without previous infection** | Reference | Reference | | |
| **Individuals with previous infection** | | | | |
| Unvaccinated | 28.9 (26.9 to 30.9) | 85.6 (82.7 to 88.0) | Reference | Reference |
| **Ad26.COV2.S** | | | | |
| First dose 0-1 wk | * | ** | * | ** |
| First dose ≥2 wk | 39.7 (37.5 to 41.8) | 91.2 (87.2 to 93.9) | 16.2 (12.4 to 19.8) | 39.5 (8.3 to 60.0) |
| Second dose 0-1 wk | 52.4 (46.9 to 57.4) | 93.3 (72.9 to 98.3) | 34.1 (26.2 to 41.1) | 55.2 (-82.7 to 89) |
| Second dose 2-9 wk | 44.8 (42.4 to 47.2) | 97.8 (94.0 to 99.2) | 22.8 (18.8 to 26.6) | 84.0 (56.6 to 94.1) |
| Second dose 10-19 wk | * | ** | * | ** |
| Second dose ≥20 wk | * | ** | * | ** |
| **BNT162b2** | | | | |
| First dose 0-1 wk | 71.0 (63.3 to 77.1) | ** | 59.0 (47.9 to 67.7) | ** |
| First dose ≥2 wk | 56.1 (54.8 to 57.5) | 90.0 (86.4 to 92.7) | 40.6 (38.1 to 42.9) | 41.9 (16.9 to 59.4) |
| Second dose 0-1 wk | 71.1 (66.8 to 74.8) | 95.5 (67.6 to 99.4) | 60.3 (54.3 to 65.5) | 72.6 (-97.3 to 96.2) |
| Second dose 2-9 wk | 63.6 (62.5 to 64.7) | 92.0 (88 to 94.7) | 51.9 (50.0 to 53.8) | 59.6 (36.6 to 74.2) |
| Second dose 10-19 wk | 50.2 (49.4 to 50.9) | 94.7 (93.4 to 95.7) | 32.8 (30.7 to 34.7) | 67.8 (57.4 to 75.6) |
| Second dose ≥20 wk | 45.7 (43.7 to 47.7) | 94.0 (91.4 to 95.9) | 26.2 (22.8 to 29.4) | 53.6 (30.2 to 69.1) |
| Booster dose 0-1 wk | 68.0 (65.8 to 70.2) | 100 (†) | 55.7 (52.3 to 58.9) | 100 (†) |
| Booster dose 2-9 wk | 68.2 (66.4 to 69.9) | 96.8 (94.1 to 98.2) | 56.4 (53.7 to 59.0) | 75.0 (53.3 to 86.7) |
| Booster dose ≥10 wk | 58.2 (45.4 to 68.1) | ** | 43.3 (25.8 to 56.6) | ** |
| **ChAdOx1** | | | | |
| First dose 0-1 wk | * | ** | * | ** |
| First dose ≥2 wk | 39.1 (36.9 to 41.2) | 88.0 (84.5 to 90.7) | 15.2 (11.3 to 18.8) | 14.3 (-16.9 to 37.2) |
| Second dose 0-1 wk | 59.7 (49.7 to 67.7) | ** | 43.6 (29.4 to 54.9) | ** |
| Second dose 2-9 wk | 45.5 (42.6 to 48.3) | 89.9 (81.9 to 94.3) | 25.5 (21 to 29.7) | 41.0 (-8.1 to 67.8) |
| Second dose 10-19 wk | 38.8 (37.7 to 39.8) | 93.9 (92.8 to 94.9) | 14.5 (11.9 to 17.1) | 57.1 (44.8 to 66.7) |
| Second dose ≥20 wk | 40.7 (39.6 to 41.7) | 94.5 (93.8 to 95.1) | 17.0 (14.4 to 19.6) | 55.4 (44.6 to 64.1) |
| Booster dose 0-1 wk | 68.8 (67.5 to 70.0) | 97.6 (96.2 to 98.5) | 56.0 (53.8 to 58.0) | 79.4 (66.8 to 87.3) |
| Booster dose 2-9 wk | 72.1 (71.4 to 72.8) | 98.1 (97.7 to 98.5) | 60.5 (59.1 to 61.9) | 84.5 (79.4 to 88.4) |
| Booster dose ≥10 wk | 50.8 (48.9 to 52.7) | 97.2 (96.2 to 98.0) | 32.6 (29.4 to 35.7) | 81.2 (72.5 to 87.1) |
| **CoronaVac** | | | | |
| First dose 0-1 wk | 51.2 (33.4 to 64.2) | ** | 30.0 (4.0 to 48.8) | ** |
| First dose ≥2 wk | 42.1 (39.8 to 44.4) | 90.4 (86.5 to 93.2) | 20.0 (16.1 to 23.6) | 40.0 (11.7 to 59.3) |
| Second dose 0-1 wk | 53.3 (43.3 to 61.4) | ** | 34.5 (20.4 to 46.1) | ** |
| Second dose 2-9 wk | 46.0 (42.6 to 49.2) | 88.4 (77.9 to 93.9) | 23.4 (18.2 to 28.3) | 34.1 (-28.9 to 66.3) |
| Second dose 10-19 wk | 31.0 (29.4 to 32.5) | 88.7 (85.4 to 91.3) | 7.3 (4.0 to 10.4) | 39.8 (16.9 to 56.4) |
| Second dose ≥20 wk | 36.2 (34.9 to 37.4) | 90.7 (89.5 to 91.8) | 12.3 (9.4 to 15.1) | 34.4 (18.3 to 47.3) |
| Booster dose 0-1 wk | 66.9 (64.7 to 69.0) | 95.0 (89.9 to 97.6) | 54.7 (51.4 to 57.8) | 67.9 (32.6 to 84.7) |
| Booster dose 2-9 wk | 73.4 (72.4 to 74.3) | 96.9 (96.0 to 97.6) | 62.7 (61.0 to 64.3) | 76.6 (68.1 to 82.8) |
| Booster dose ≥10 wk | 54.6 (53.7 to 55.5) | 96.7 (96.2 to 97.1) | 37.9 (35.8 to 40.0) | 75.7 (69.6 to 80.7) |

*to ensure reasonable precision estimates are given if there are at least 20 cases or 1000 controls - Symptomatic infection

**to ensure reasonable precision estimates are shown if there are at least 10 cases or 500 controls - Severe outcomes

† No cases

Supplementary Table 4: Protection conferred by the previous infection, stratified by time since the first SARS-CoV-2 infection

| **Status** | **Protection% (95% CI)** | |
|---|---|---|
| | Symptomatic Infection | Severe Outcomes |
| **Unvaccinated without previous infection** | Reference | Reference |
| **Unvaccinated with previous infection** | | |
| 3-5 months ago | 52.8 (48.3 to 56.8) | 84.5 (73.1 to 91.1) |
| 6-12 months ago | 32.7 (30.2 to 35.2) | 89.5 (86 to 92.2) |
| > 1 year ago | 14.7 (10.8 to 18.5) | 80.3 (74.4 to 84.8) |

Supplementary Table 5: Characteristics of individuals tested for SARS-CoV-2 in Brazil during the Omicron period, stratified by case/control – Matched Design (1:2 ratio with replacement)

| Characteristic | Cases, N = 423,068 | Controls, N = 816,294 | Total, N = 1,239,362 |
|---|---|---|---|
| **No. individuals** | 419,617 (99.2%) | 272,133 (33.3%) | 691,750 (55.8%) |
| **Age median(IQR)-years** | 37 (29, 47) | 37 (28, 46) | 37 (28, 46) |
| **Age Group** | | | |
| 18-64 | 404,764 (95.7%) | 784,509 (96.1%) | 1,189,273 (96.0%) |
| ≥ 65 | 18,304 (4.3%) | 31,785 (3.9%) | 50,089 (4.0%) |
| **Sex-Female** | 239,032 (56.5%) | 462,948 (56.7%) | 701,980 (56.6%) |
| **Race** | | | |
| White | 217,112 (51.3%) | 425,475 (52.1%) | 642,587 (51.8%) |
| Black | 16,695 (3.9%) | 36,465 (4.5%) | 53,160 (4.3%) |
| Asian | 7,994 (1.9%) | 16,278 (2.0%) | 24,272 (2.0%) |
| Mixed | 134,939 (31.9%) | 271,588 (33.3%) | 406,527 (32.8%) |
| Indigenous | 470 (0.1%) | 1,385 (0.2%) | 1,855 (0.1%) |
| (Missing) | 45,858 (10.8%) | 65,103 (8.0%) | 110,961 (9.0%) |
| **Previous SARS-CoV-2 Infection** | | | |
| No | 162,474 (38.4%) | 200,382 (24.5%) | 362,856 (29.3%) |
| 90-179d | 12,489 (3.0%) | 43,182 (5.3%) | 55,671 (4.5%) |
| 180-365d | 133,061 (31.5%) | 333,352 (40.8%) | 466,413 (37.6%) |
| 366+d | 115,044 (27.2%) | 239,378 (29.3%) | 354,422 (28.6%) |
| **Test type** | | | |
| Lateral-flow | 330,769 (78.2%) | 680,104 (83.3%) | 1,010,873 (81.6%) |
| RT-PCR | 92,299 (21.8%) | 136,190 (16.7%) | 228,489 (18.4%) |
| **Residence in capital state** | 89,789 (21.2%) | 179,265 (22.0%) | 269,054 (21.7%) |
| **Macroregion** | | | |
| North | 26,898 (6.4%) | 50,949 (6.2%) | 77,847 (6.3%) |
| Northeast | 59,190 (14.0%) | 112,184 (13.7%) | 171,374 (13.8%) |
| Southeast | 174,323 (41.2%) | 337,873 (41.4%) | 512,196 (41.3%) |
| South | 121,441 (28.7%) | 236,565 (29.0%) | 358,006 (28.9%) |
| Central-west | 41,216 (9.7%) | 78,723 (9.6%) | 119,939 (9.7%) |
| **Municipality deprivation quintile** | | | |
| 1 (Least) | 154,142 (36.4%) | 305,495 (37.4%) | 459,637 (37.1%) |
| 2 | 83,480 (19.7%) | 163,022 (20.0%) | 246,502 (19.9%) |
| 3 | 81,409 (19.2%) | 156,054 (19.1%) | 237,463 (19.2%) |
| 4 | 69,540 (16.4%) | 130,522 (16.0%) | 200,062 (16.1%) |
| 5 (Most) | 34,395 (8.1%) | 61,037 (7.5%) | 95,432 (7.7%) |
| (Missing) | 102 (0.0%) | 164 (0.0%) | 266 (0.0%) |
| **Diabetes Mellitus** | 12,862 (3.0%) | 30,592 (3.7%) | 43,454 (3.5%) |
| **Obesity** | 6,436 (1.5%) | 15,314 (1.9%) | 21,750 (1.8%) |
| **Immunosuppression** | 3,179 (0.8%) | 6,981 (0.9%) | 10,160 (0.8%) |
| **Chronic respiratory disease** | 13,500 (3.2%) | 32,587 (4.0%) | 46,087 (3.7%) |
| **Chronic cardiac disease** | 24,444 (5.8%) | 57,360 (7.0%) | 81,804 (6.6%) |
| **Chronic Kidney disease** | 1,624 (0.4%) | 3,162 (0.4%) | 4,786 (0.4%) |
| **Number of medical comorbidities** | | | |
| 0 | 373,894 (88.4%) | 702,106 (86.0%) | 1,076,000 (86.8%) |
| 1 | 38,568 (9.1%) | 88,202 (10.8%) | 126,770 (10.2%) |
| 2 | 8,649 (2.0%) | 21,037 (2.6%) | 29,686 (2.4%) |
| ≥3 | 1,957 (0.5%) | 4,949 (0.6%) | 6,906 (0.6%) |
| **Hospitalization** | 7,303 (1.7%) | 7,608 (0.9%) | 14,911 (1.2%) |
| **Death** | 2,406 (0.6%) | 1,604 (0.2%) | 4,010 (0.3%) |
| **Severe outcomes** | 7,746 (1.8%) | 8,035 (1.0%) | 15,781 (1.3%) |

Supplementary Table 6: Vaccination Status of individuals tested for SARS-CoV-2 in Brazil, according to the test result and severity – Matched Design (1:2 ratio with replacement)

| Status | Controls, N = 816,294 | Symptomatic Infection, N = 415,322 | Severe Outcomes, N = 7,746 |
|---|---|---|---|
| **Unvaccinated without previous infection** | 200,382 (55.2%) | 156,128 (43.0%) | 6,346 (1.7%) |
| **Individuals with previous infection** | | | |
| Unvaccinated | 20,163 (63.9%) | 11,303 (35.8%) | 108 (0.3%) |
| **Ad26.COV2.S** | | | |
| First dose 0-1 wk | 40 (76.9%) | 12 (23.1%) | 0 (0.0%) |
| First dose ≥2 wk | 13,117 (66.6%) | 6,540 (33.2%) | 25 (0.1%) |
| Second dose 0-1 wk | 1,457 (71.0%) | 594 (28.9%) | 2 (0.1%) |
| Second dose 2-9 wk | 8,901 (68.3%) | 4,136 (31.7%) | 3 (0.0%) |
| Second dose 10-19 wk | 297 (65.7%) | 154 (34.1%) | 1 (0.2%) |
| Second dose ≥20 wk | 0 (0.0%) | 6 (100.0%) | 0 (0.0%) |
| **BNT162b2** | | | |
| First dose 0-1 wk | 421 (80.2%) | 103 (19.6%) | 1 (0.2%) |
| First dose ≥2 wk | 20,478 (73.7%) | 7,273 (26.2%) | 32 (0.1%) |
| Second dose 0-1 wk | 1,235 (81.0%) | 289 (19.0%) | 1 (0.1%) |
| Second dose 2-9 wk | 24,765 (77.9%) | 7,024 (22.1%) | 19 (0.1%) |
| Second dose 10-19 wk | 99,861 (71.2%) | 40,356 (28.8%) | 81 (0.1%) |
| Second dose ≥20 wk | 11,964 (69.3%) | 5,272 (30.5%) | 24 (0.1%) |
| Booster dose 0-1 wk | 4,731 (79.0%) | 1,259 (21.0%) | 0 (0.0%) |
| Booster dose 2-9 wk | 6,803 (78.6%) | 1,848 (21.3%) | 9 (0.1%) |
| Booster dose ≥10 wk | 148 (68.2%) | 69 (31.8%) | 0 (0.0%) |
| **ChAdOx1** | | | |
| First dose 0-1 wk | 37 (71.2%) | 15 (28.8%) | 0 (0.0%) |
| First dose ≥2 wk | 12,768 (67.2%) | 6,180 (32.5%) | 53 (0.3%) |
| Second dose 0-1 wk | 353 (73.7%) | 126 (26.3%) | 0 (0.0%) |
| Second dose 2-9 wk | 6,298 (70.0%) | 2,687 (29.9%) | 9 (0.1%) |
| Second dose 10-19 wk | 78,941 (66.8%) | 39,199 (33.1%) | 111 (0.1%) |
| Second dose ≥20 wk | 62,368 (66.3%) | 31,485 (33.5%) | 246 (0.3%) |
| Booster dose 0-1 wk | 13,923 (78.9%) | 3,707 (21.0%) | 15 (0.1%) |
| Booster dose 2-9 wk | 42,156 (80.3%) | 10,298 (19.6%) | 61 (0.1%) |
| Booster dose ≥10 wk | 11,215 (71.0%) | 4,559 (28.9%) | 26 (0.2%) |
| **CoronaVac** | | | |
| First dose 0-1 wk | 165 (70.5%) | 69 (29.5%) | 0 (0.0%) |
| First dose ≥2 wk | 11,220 (69.3%) | 4,942 (30.5%) | 29 (0.2%) |
| Second dose 0-1 wk | 448 (70.6%) | 187 (29.4%) | 0 (0.0%) |
| Second dose 2-9 wk | 4,704 (70.4%) | 1,967 (29.4%) | 10 (0.1%) |
| Second dose 10-19 wk | 36,141 (65.4%) | 19,063 (34.5%) | 56 (0.1%) |
| Second dose ≥20 wk | 45,561 (65.7%) | 23,571 (34.0%) | 238 (0.3%) |
| Booster dose 0-1 wk | 5,186 (79.1%) | 1,362 (20.8%) | 8 (0.1%) |
| Booster dose 2-9 wk | 19,688 (81.1%) | 4,542 (18.7%) | 46 (0.2%) |
| Booster dose ≥10 wk | 50,359 (72.4%) | 18,997 (27.3%) | 186 (0.3%) |

Supplementary Table 7: Effectiveness estimates derived from the matched design

| **Status** | **VE% (95% CI)** | | | |
|---|---|---|---|---|
| | **Symptomatic Infection** | **Severe Outcomes** | **Symptomatic Infection** | **Severe Outcomes** |
| **Unvaccinated without previous infection** | Reference | Reference | | |
| **Individuals with previous infection** | | | | |
| **Unvaccinated** | 31.7 (30 to 33.4) | 81.9 (76.2 to 86.3) | Reference | Reference |
| **Ad26.COV2.S** | | | | |
| First dose 0-1 wk | * | ** | * | ** |
| First dose ≥2 wk | 41.2 (39.4 to 43) | 89.4 (82.1 to 93.8) | 16.9 (13.2 to 20.5) | 45.4 (-19.6 to 75.1) |
| Second dose 0-1 wk | 53.3 (48.5 to 57.6) | 85.4 (16.9 to 97.4) | 32.8 (25 to 39.8) | 37.9 (-659.4 to 94.9) |
| Second dose 2-9 wk | 47.2 (45.2 to 49.2) | 97.5 (91.3 to 99.3) | 24.5 (20.6 to 28.2) | 92.3 (64.7 to 98.3) |
| Second dose 10-19 wk | 46.5 (34.7 to 56.2) | ** | 16.2 (-4.7 to 33) | ** |
| Second dose ≥20 wk | * | ** | * | |
| **BNT162b2** | | | | |
| First dose 0-1 wk | 70.7 (63.6 to 76.4) | ** | 58.4 (46.7 to 67.5) | ** |
| First dose ≥2 wk | 57.1 (55.8 to 58.3) | 88.9 (82.2 to 93.1) | 39.2 (36.7 to 41.6) | 60 (15.4 to 81.1) |
| Second dose 0-1 wk | 72.2 (68.3 to 75.6) | 87.4 (-44 to 98.9) | 60.3 (54.1 to 65.6) | ** |
| Second dose 2-9 wk | 66.5 (65.5 to 67.5) | 90.9 (84 to 94.8) | 54.1 (52.1 to 55.9) | 53.6 (-6.4 to 79.8) |
| Second dose 10-19 wk | 53.1 (52.5 to 53.8) | 93.4 (91.2 to 95) | 33.9 (31.8 to 35.8) | 72.9 (52.6 to 84.5) |
| Second dose ≥20 wk | 50.1 (48.4 to 51.8) | 91.1 (85.3 to 94.6) | 30.6 (27.3 to 33.7) | 55.1 (-1.9 to 80.2) |
| Booster dose 0-1 wk | 70.2 (68.3 to 72) | 100 (†) | 56.1 (52.8 to 59.2) | 100 (†) |
| Booster dose 2-9 wk | 70 (68.4 to 71.6) | 95.7 (90.6 to 98) | 58.1 (55.3 to 60.6) | 85.2 (55.7 to 95.1) |
| Booster dose ≥10 wk | 51 (34.3 to 63.5) | ** | 29.8 (3.3 to 49) | ** |
| **ChAdOx1** | | | | |
| First dose 0-1 wk | * | ** | * | ** |
| First dose ≥2 wk | 41.6 (39.7 to 43.4) | 85.6 (78.6 to 90.3) | 17.1 (13.3 to 20.6) | 31.4 (-38.3 to 66) |
| Second dose 0-1 wk | 57.7 (48.1 to 65.6) | ** | 42.5 (27.7 to 54.2) | ** |
| Second dose 2-9 wk | 49 (46.6 to 51.3) | 90.2 (77.4 to 95.8) | 27.2 (22.9 to 31.3) | 67.5 (-7.9 to 90.2) |
| Second dose 10-19 wk | 42.3 (41.5 to 43.2) | 92.4 (90.3 to 94.1) | 16.8 (14.2 to 19.3) | 70.4 (48.6 to 83) |
| Second dose ≥20 wk | 41.4 (40.5 to 42.3) | 92.7 (91.2 to 93.9) | 15.9 (13.2 to 18.5) | 63.2 (39 to 77.8) |
| Booster dose 0-1 wk | 69.7 (68.6 to 70.9) | 96.5 (93.8 to 98.1) | 56 (53.8 to 58) | 82.9 (60.8 to 92.6) |
| Booster dose 2-9 wk | 72.9 (72.2 to 73.5) | 97.5 (96.6 to 98.1) | 61.3 (59.9 to 62.7) | 89.7 (81.5 to 94.3) |
| Booster dose ≥10 wk | 54.8 (53.1 to 56.4) | 96.6 (94.7 to 97.9) | 36.4 (33.3 to 39.4) | 86.3 (71.6 to 93.4) |
| **CoronaVac** | | | | |
| First dose 0-1 wk | 51.8 (35.9 to 63.7) | ** | 25.8 (-1.5 to 45.8) | ** |
| First dose ≥2 wk | 46.3 (44.4 to 48.2) | 91.6 (86 to 95) | 24.2 (20.6 to 27.7) | 69.5 (35.9 to 85.4) |
| Second dose 0-1 wk | 52 (43 to 59.6) | ** | 26.3 (10.6 to 39.3) | ** |
| Second dose 2-9 wk | 49.3 (46.5 to 52) | 78.4 (48.2 to 91) | 27.3 (22.3 to 31.9) | 21.4 (-148.4 to 75.1) |
| Second dose 10-19 wk | 36.5 (35.2 to 37.8) | 89.2 (84.6 to 92.4) | 10.8 (7.7 to 13.8) | 66.4 (37.6 to 81.9) |
| Second dose ≥20 wk | 39.4 (38.3 to 40.5) | 89.3 (87.1 to 91.1) | 14.3 (11.4 to 17) | 46.8 (11.4 to 68) |
| Booster dose 0-1 wk | 69.9 (68 to 71.6) | 91.5 (80.2 to 96.4) | 57.4 (54.3 to 60.4) | 65 (-11.9 to 89.1) |
| Booster dose 2-9 wk | 74 (73.1 to 74.8) | 95.9 (94.1 to 97.1) | 62.9 (61.2 to 64.5) | 81 (64 to 90) |
| Booster dose ≥10 wk | 57.2 (56.4 to 58) | 96.5 (95.8 to 97.2) | 41.1 (39.1 to 43.1) | 84.5 (74 to 90.8) |

*to ensure reasonable precision estimates are given if there are at least 20 cases or 1000 controls - Symptomatic infection

**to ensure reasonable precision estimates are shown if there are at least 10 cases or 500 controls - Severe outcomes

† No cases
